# Beyond Malodor: A Multinational Patient-Reported Study of the Diagnostic, Psychosocial, and Cognitive Burden of Trimethylaminuria

**DOI:** 10.64898/2026.09.08.26362298

**Authors:** Luz Ayda Gomez Lopez, Sang Sun Yoon, Vanessa Munoz Espana, Carolina Valero Walteros

**Affiliations:** Fundación Universitaria del Área Andina, Bogotá, Colombia; Department of Microbiology and Immunology, Institute for Immunology and Immunological Diseases, Yonsei University College of Medicine, Seoul, Republic of Korea; BioMe Inc., Seoul, Republic of Korea; School of Medicine, Faculty of Health Sciences, Universidad de Nariño, Colombia; International Foundation for Metabolic Diseases, Bogota, Colombia

**Author notes:** Correspondence to Luz Ayda Gomez Lopez, Address: Fundación Universitaria del Área Andina, Carrera 14 A No. 70 A - 34, Bogotá, D.C., Colombia. LAGL, SSY and VME contributed equally to this work as co-first authors.

**Keywords:** Trimethylaminuria, TMAU, rare disease, patient-reported outcomes, stigma, cognitive symptoms, cognitive dysfunction, quality of life

## Abstract

**Background:** Trimethylaminuria (TMAU) is a rare metabolic disorder characterized by malodor resulting from impaired metabolism of trimethylamine, but its broader impact on patients’ lives remains insufficiently characterized. This study aimed to assess the diagnostic, psychosocial, educational, occupational, and treatment-related burden of TMAU across a multinational patient population and to explore cognitive manifestations identified during the study.

**Methods:** We conducted a multinational, cross-sectional, patient-reported survey using an anonymous online questionnaire. Of 450 submissions, 401 participants provided valid authorization for analysis. The survey assessed demographic characteristics, symptom and diagnostic experiences, educational and occupational consequences, psychosocial burden, and perceptions of current management. Cognitive complaints emerging during the initial study prompted a follow-up survey focused on cognitive symptoms and their functional impact, completed by 306 participants from the original cohort. The association between cognitive symptom frequency and severity was assessed using Spearman rank correlation.

**Results:** Participants represented more than 40 countries, and 61.1% were female. A substantial diagnostic gap was observed, with 65.8% reporting no official diagnosis or clear medical explanation of probable TMAU. Social and psychological consequences were prominent, including physical rejection or social exclusion (78.8%), bullying or derogatory treatment (61.1%), and substantial disruption of educational and occupational participation. Most participants (84.3%) considered currently available treatment options insufficient. In the cognitive follow-up survey, 68.0% reported cognitive difficulties at least several times per week, 58.8% reported moderate or severe symptoms, and 66.7% reported at least moderate interference with daily activities, work, or studies. Greater cognitive symptom frequency was significantly associated with greater severity (Spearman’s ρ = 0.451, p < 0.001).

**Conclusions:** TMAU is associated with a multidimensional patient-reported burden extending beyond malodor to psychosocial, educational, occupational, and previously underrecognized cognitive manifestations. These findings support earlier recognition, multidisciplinary care, and development of mechanism-based therapies evaluated using patient-centered outcomes that include daily functioning, cognitive symptoms, and quality of life.

## Background

Trimethylaminuria (TMAU) is a metabolic disorder characterized by the accumulation of trimethylamine (TMA), a volatile amine with a strong odor commonly described as resembling decaying fish. Primary TMAU is most often caused by pathogenic variants in the flavin-containing monooxygenase 3 (FMO3) gene, which impair the hepatic conversion of odorous TMA into the largely odorless metabolite trimethylamine N-oxide (TMAO). Secondary forms may develop in the absence of pathogenic FMO3 variants and have been associated with alterations in gut microbial TMA production, impaired hepatic or renal function, and other metabolic or hormonal factors [1]. The severity of malodor may fluctuate over time and can worsen during periods of physiological or hormonal change. In women, symptoms may first become apparent or intensify during puberty, adolescence, menstruation, hormonal disturbances, or oral contraceptive use [2, 3]. Episodes of fever, strenuous exercise, and emotional stress may further exacerbate odor by increasing perspiration and the release of volatile TMA. Because TMA is excreted through sweat, urine, breath, and other bodily secretions, it can disperse into the surrounding air and produce a persistent and socially perceptible body odor.

The population prevalence of TMAU remains uncertain, and reliable population-based incidence data are not currently available. Historical phenotyping studies estimated the frequency of reduced TMA N-oxidation at approximately 0.5∼1.0% in White British populations, with higher frequencies reported in some other populations, including 1.7% in Jordan, 3.8% in Ecuador, and 11.0% in New Guinea [4, 5]. However, these estimates reflect carrier or reduced metabolic-phenotype frequencies and should not be interpreted as the prevalence of clinically manifest TMAU. The true disease burden is difficult to determine because of variable phenotypic expression, intermittent or fluctuating symptoms, limited diagnostic testing, and frequent clinical underrecognition. Consequently, formally diagnosed cases are likely to represent only a fraction of the affected population [4, 6].

Although rare, TMAU can cause substantial psychosocial impairment because of its stigmatizing odor and the absence of definitive treatment. Patients often report repeated consultations from childhood or early adulthood without receiving an accurate diagnosis. In the absence of adequate clinical guidance, many turn to online communities, restrictive diets, supplements, or empirically selected interventions, often with inconsistent benefit [7]. The burden may be especially severe when symptoms emerge during childhood or adolescence, when self-concept, identity, and peer relationships are developing. Social rejection, ridicule, and exclusion may contribute to reduced self-esteem, anxiety, and social withdrawal, highlighting the need for age-appropriate psychosocial support [8].

Systematic patient-reported evidence remains limited. A recent UK survey of 47 adults and caregivers documented diagnostic barriers, social exclusion, discrimination, and psychological distress, but was limited by its small, single-country sample and inclusion of individuals without confirmed TMAU [9]. The present study addresses these limitations through a multinational cohort of more than 400 individuals with self-reported or clinically diagnosed TMAU. By assessing diagnostic pathways, educational and social experiences, bullying, and psychosocial outcomes, it provides the largest cross-cultural patient-reported assessment of TMAU to date and a foundation for earlier recognition, psychosocial support, and safeguarding policies in clinical and educational settings.

## Methods

### Study design and participants

We conducted a multinational, cross-sectional, patient-reported survey using KoboToolbox to assess the burden of trimethylaminuria (TMAU). Participants were recruited primarily through international TMAU patient communities and online networks and included individuals who self-reported TMAU or had received a clinical diagnosis or medical explanation consistent with the condition. A total of 450 submissions were received. After exclusion of participants who declined or did not complete the applicable data-use authorization process, 401 participants were included in the primary analysis. Adults provided electronic informed consent; minors participated with parental or legal-guardian consent and their own assent.

### Ethics

Participation was voluntary, and all responses were analyzed and reported in aggregate form. The informed-consent process was developed and reviewed by the study team and approved by the Latin American Patients’ Academy (ALAPA).

### Survey measures

The questionnaire assessed demographic characteristics, symptom onset and pubertal changes, diagnostic experiences, educational and occupational consequences, psychological and social burden, sleep disturbance, perceived adequacy of current management, and willingness to permit anonymized data to support scientific or regulatory purposes. Questions were structured as single- or multiple-response items as appropriate. Several retrospective age-at-event questions permitted more than one response because participants could be uncertain about the timing of past events. For multiple-response items, percentages were calculated relative to the analytical cohort and therefore could cumulatively exceed 100%.

Participants were also given opportunities to describe their experiences in their own words through open-ended responses. These qualitative comments were used to provide additional context for the quantitative findings, and representative anonymized responses are presented in the Supplementary Information.

During the initial study, cognitive complaints emerged as a recurring concern. A follow-up questionnaire was therefore administered to assess patient-reported “brain fog,” including its onset, frequency, severity, affected cognitive domains, and interference with daily activities, work, or studies. This follow-up survey was completed by 306 participants, all of whom were members of the original cohort.

### Statistical analysis

Categorical data were summarized as numbers and percentages. For the cognitive analysis, we examined whether more frequent cognitive symptoms were associated with greater symptom severity using Spearman’s rank correlation. Participants who reported no cognitive symptoms or had missing frequency or severity data were excluded, leaving 282 participants for this analysis. A two-sided p value < 0.05 was considered statistically significant. All other analyses were descriptive.

## Results

### Study cohort and demographic characteristics

A total of 450 survey submissions were recorded. Of these, 391 adults provided authorization for anonymized use of their data, and 10 minors participated with legal-guardian authorization and their own assent, yielding an analytical cohort of 401 participants. Nine adults declined data-use authorization, while 40 additional submissions did not complete or satisfy the applicable consent-validation process and were excluded from analysis. All demographic summaries in Table 1 therefore use N = 401 as the denominator.

**Table 1.**
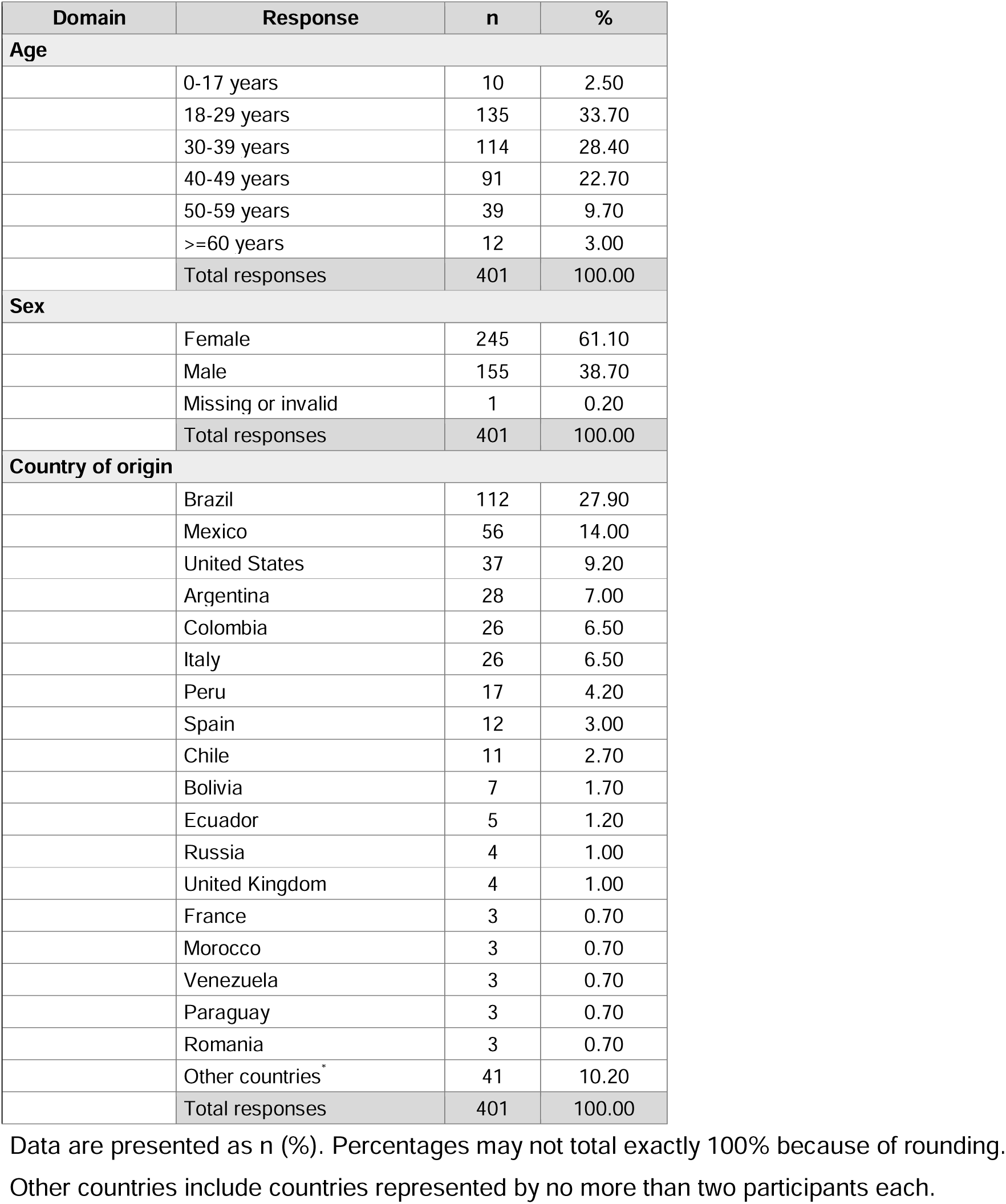
Demographic and geographic characteristics of the study cohort (N = 401)

The cohort was predominantly adult. Participants aged 18-29 years comprised the largest age group (n = 135, 33.7%), followed by those aged 30-39 years (n = 114, 28.4%) and 40-49 years (n = 91, 22.7%). Collectively, 340 participants (84.8%) were between 18 and 49 years of age. Participants aged 50-59 years accounted for 39 responses (9.7%), 12 participants (3.0%) were 60 years of age or older, and 10 (2.5%) were younger than 18 years. Although the survey included items addressing childhood and adolescent experiences, the analytical cohort was predominantly adult; therefore, earlier-life experiences were reported retrospectively (Tables 1–4).

**Table 2.**
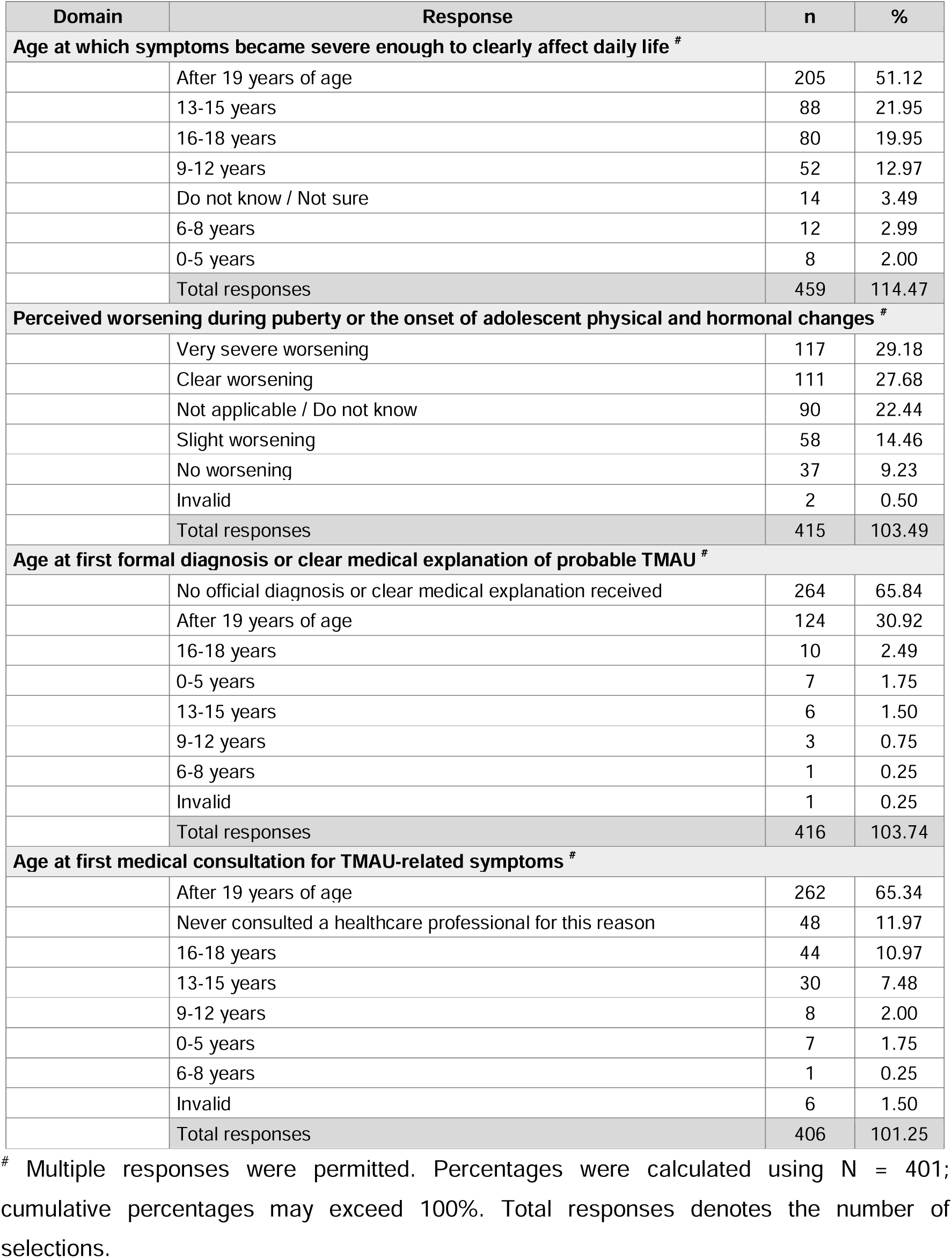
Symptom onset, pubertal worsening, and diagnostic pathway (N = 401)

**Table 3.** Educational and professional consequences of TMAU (N = 401)

| Domain | Response | n | % |
| --- | --- | --- | --- |
| **School absence before 18 years of age due to symptoms or fear related to body odor** |  |  |  |
|  | No school absence | 207 | 51.62 |
|  | Prolonged absence, school dropout, or homeschooling | 59 | 14.71 |
|  | More than 10 days per semester | 47 | 11.72 |
|  | 1-2 days per semester | 34 | 8.48 |
|  | 3-5 days per semester | 24 | 5.99 |
|  | 6-10 days per semester | 29 | 7.23 |
|  | Invalid | 1 | 0.25 |
|  | Total responses | 401 | 100.00 |
| **Impact on educational trajectory before 18 years of age <sup>#</sup>** |  |  |  |
|  | No change in educational trajectory | 161 | 40.15 |
|  | Reduced active participation in class | 124 | 30.92 |
|  | Withdrawal from recreational, sports, or extracurricular activities | 122 | 30.42 |
|  | Abandonment or modification of future higher-education or vocational plans | 98 | 24.44 |
|  | Permanent withdrawal from school or prolonged disruption of education | 68 | 16.96 |
|  | Transfer to a different educational institution | 33 | 8.23 |
|  | Transition to virtual/online classes or homeschooling | 24 | 5.99 |
|  | Total responses | 630 | 157.11 |
| **Impact before 18 years of age on future academic and professional plans or goals <sup>#</sup>** |  |  |  |
|  | Relinquished aspirations for leadership positions or public-facing professions | 185 | 46.13 |
|  | Developed a preference for isolated or remote work environments | 174 | 43.39 |
|  | Avoided professional networking or career connections due to fear | 160 | 39.90 |
|  | Felt hopeless about future employment prospects | 149 | 37.16 |
|  | No effect on future professional planning | 68 | 16.96 |
|  | Prefer not to answer | 18 | 4.49 |
|  | Total responses | 754 | 188.03 |

2 The school-absence item was treated as a single-response item. <sup>#</sup> The educational-trajectory 3 and future-planning items allowed multiple responses; percentages are based on N = 401 4 and may exceed 100%. Total responses denotes the number of selections.

**Table 4.** Psychological, social, and functional burden associated with TMAU (N = 401)

| Domain | Response | n | % |
| --- | --- | --- | --- |
| **Psychological and psychiatric experiences before 18 years of age <sup>#</sup>** |  |  |  |
|  | Persistent anxiety symptoms or panic attacks | 191 | 47.63 |
|  | Clinical depression or profound feelings of hopelessness | 174 | 43.39 |
|  | Suicidal ideation | 164 | 40.90 |
|  | None of the above | 107 | 26.68 |
|  | Self-harm thoughts or behaviors | 80 | 19.95 |
|  | Psychotherapy, counseling, or consultation with a mental-health professional | 66 | 16.46 |
|  | Prescription of anxiolytic or antidepressant medication | 62 | 15.46 |
|  | Prefer not to answer | 22 | 5.49 |
|  | Crisis hotline use, emergency-room visit, or urgent psychiatric intervention | 19 | 4.74 |
|  | Total responses | 885 | 220.70 |
| **Comparison of psychosocial impact before 18 years of age and in adulthood <sup>#</sup>** |  |  |  |
|  | Impact in adulthood was much more severe | 232 | 57.86 |
|  | Impact was very similar at both life stages | 76 | 18.95 |
|  | Impact before 18 years of age was much more severe | 56 | 13.97 |
|  | Impact in adulthood was slightly more severe | 41 | 10.22 |
|  | Not applicable (minor or not yet fully adult) | 25 | 6.23 |
|  | Impact before 18 years of age was slightly more severe | 11 | 2.74 |
|  | Total responses | 441 | 109.98 |
| **Adverse family, school, workplace, or social experiences despite rigorous hygiene efforts <sup>#</sup>** |  |  |  |
|  | Direct physical rejection or social exclusion | 316 | 78.80 |
|  | Unprompted comments from family members, colleagues, supervisors, or others | 299 | 74.56 |
|  | Severe disruption of daily routines to reduce the risk of rejection | 284 | 70.82 |
|  | Bullying, harassment, mockery, or derogatory remarks | 245 | 61.10 |
|  | Formal or informal institutional complaints or sanctions | 114 | 28.43 |
|  | * None of the above; environment was fully respectful | 7 | 1.75 |
|  | Total responses | 1,265 | 315.46 |
| **Sleep difficulties caused by condition-related concern, stress, or anxiety** |  |  |  |
|  | Frequently | 147 | 36.66 |
|  | Sometimes | 114 | 28.43 |
|  | Always or almost always | 58 | 14.46 |
|  | Rarely | 56 | 13.97 |
|  | Never | 18 | 4.49 |
|  | Invalid | 8 | 2.00 |
|  | Total responses | 401 | 100.00 |

<sup>#</sup> Multiple responses were permitted. Percentages were calculated using N = 401; cumulative percentages may exceed 100%. Total responses denotes the number of selections. \* "None of the above" option was mutually exclusive.

Female participants were more common than male participants, with 245 females (61.1%) and 155 males (38.7%); one record (0.2%) had missing or invalid sex information. Geographic representation was broad, with respondents from more than 40 countries. The largest groups were from Brazil (n = 112, 27.9%), Mexico (n = 56, 14.0%), the United States (n = 37, 9.2%), Argentina (n = 28, 7.0%), Colombia (n = 26, 6.5%), and Italy (n = 26, 6.5%). These six countries accounted for 285 of 401 participants (71.1%), while the remainder were distributed across numerous countries in Latin America, Europe, North America, and other regions (Table 1).

### Symptom onset, pubertal worsening, and diagnostic pathway

Symptoms severe enough to affect daily life were most frequently reported after 19 years of age (205 selections, 51.1% relative to N = 401), but substantial numbers indicated onset during adolescence, including 13-15 years (n = 88, 21.9%) and 16-18 years (n = 80, 20.0%); 52 selections (13.0%) indicated ages 9-12 years (Table 2). Perceived worsening around puberty was also prominent, with very severe worsening selected 117 times (29.2%), clear worsening 111 times (27.7%), and slight worsening 58 times (14.5%).

Diagnostic recognition was limited: no official diagnosis or clear medical explanation of probable TMAU was selected 264 times (65.8%), while diagnosis or a clear explanation after age 19 was selected 124 times (30.9%). First consultation after age 19 was selected 262 times (65.3%), and 48 selections (12.0%) indicated that no healthcare professional had ever been consulted for these symptoms. Although multiple selections were permitted for these retrospective age-at-event items, response totals only modestly exceeded the cohort size (416 selections for age at diagnosis or medical explanation and 406 for age at first consultation, vs. N = 401), indicating that most participants selected a single age category while a small minority selected more than one. Percentages for these items therefore represent selection frequencies and should not be interpreted as mutually exclusive participant proportions.

### Educational and professional consequences

Before 18 years of age, 193 participants (48.1%) reported some degree of school absence related to symptoms or fear of body odor, including 59 (14.7%) who reported prolonged absence, school dropout, or homeschooling and 47 (11.7%) who missed more than 10 days per semester (Table 3).

Beyond attendance, reduced classroom participation (n = 124, 30.9%), withdrawal from recreational, sports, or extracurricular activities (n = 122, 30.4%), and abandonment or modification of higher-education or vocational plans (n = 98, 24.4%) were commonly selected. Future professional planning was also affected: 185 participants (46.1%) relinquished aspirations for leadership or public-facing professions, 174 (43.4%) preferred isolated or remote work, 160 (39.9%) avoided professional networking, and 149 (37.2%) reported hopelessness about future employment. Educational-trajectory and future-planning items allowed multiple responses, so these categories overlap.

### Psychological, social, and functional burden

Psychological burden before 18 years of age was substantial (Table 4). Persistent anxiety symptoms or panic attacks were selected by 191 participants (47.6%), clinical depression or profound hopelessness by 174 (43.4%), suicidal ideation by 164 (40.9%), and self-harm thoughts or behaviors by 80 (20.0%).

Adverse interpersonal and institutional experiences were also common despite rigorous hygiene efforts. Direct physical rejection or social exclusion was selected by 316 participants (78.8%), unsolicited comments by family members, colleagues, supervisors, or others by 299 (74.6%), severe disruption of daily routines to reduce the risk of rejection by 284 (70.8%), and bullying, harassment, mockery, or derogatory remarks by 245 (61.1%). Formal or informal complaints or sanctions were reported by 114 participants (28.4%).

Sleep disturbance related to condition-associated concern, stress, or anxiety was reported frequently or always/almost always by 205 participants (51.1%), and sometimes by another 114 (28.4%). When participants compared life stages, the most frequently selected response was that psychosocial impact was much more severe in adulthood (232 selections, 57.9%). Multiple-response items in Table 4 are reported as selection frequencies where applicable.

### Qualitative patient narratives

To complement the quantitative survey findings, 29 open-ended participant narratives are provided in Additional file 1. These accounts illustrated recurrent experiences of stigma and bullying, social withdrawal and avoidance of public spaces, disruption of education and employment, strain on family and interpersonal relationships, and substantial psychological distress, including anxiety, depressive symptoms, and suicidal ideation. Participants also described major effects on daily functioning, including avoidance of public transportation, travel, school attendance, and in-person work. These narratives provide additional context for the multidimensional burden identified in the quantitative analyses.

### Perceived adequacy of management and willingness to support regulatory review

Current management was widely perceived as inadequate (Table 5). Overall, 286 participants (71.3%) agreed or strongly agreed that current medical recommendations, restrictive diets, and hygiene products were insufficient to provide an acceptable and dignified quality of life. In parallel, 338 (84.3%) rated available medical treatment options as insufficient because effective options were absent or existing approaches did not work, whereas only two participants (0.5%) considered accessible and highly effective treatment options to be available.

**Table 5.** Perceived adequacy of current management and willingness to support regulatory review (N = 401)

| Domain | Response | n | % |
| --- | --- | --- | --- |
| **Agreement that current medical recommendations, restrictive diets, and hygiene products are insufficient for an acceptable and dignified quality of life** |  |  |  |
|  | Strongly agree | 196 | 48.88 |
|  | Agree | 90 | 22.44 |
|  | Neutral | 52 | 12.97 |
|  | Disagree | 26 | 6.48 |
|  | Strongly disagree | 23 | 5.74 |
|  | Invalid | 14 | 3.49 |
|  | Total responses | 401 | 100.00 |
| **Perceived availability of effective medical treatment options** |  |  |  |
|  | Insufficient: no effective options exist or existing options do not work | 338 | 84.29 |
|  | Moderately sufficient: effectiveness or accessibility is limited | 57 | 14.21 |
|  | Sufficient: accessible and highly effective treatments exist | 2 | 0.50 |
|  | Invalid | 4 | 1.00 |
|  | Total responses | 401 | 100.00 |
| **Willingness to allow fully anonymized experience to be used in regulatory patient testimony or case dossiers** |  |  |  |
|  | Yes | 363 | 90.52 |
|  | No; retain data for statistical analysis and scientific publication only | 35 | 8.73 |
|  | Invalid | 3 | 0.75 |
|  | Total responses | 401 | 100.00 |

Data are presented as n (%). Percentages were calculated using N = 401 and may not total exactly 100% because of rounding.

A large majority (n = 363, 90.5%) were willing to allow their fully anonymized experiences to be used in regulatory patient testimony or case dossiers; 35 (8.7%) preferred use to remain limited to statistical analysis and scientific publication.

### Cognitive manifestations and functional impact of brain fog

During the course of the initial study, we noted that a number of participants spontaneously reported cognitive complaints, including difficulties with memory, concentration, and mental clarity. To characterize these manifestations more systematically, we subsequently conducted a follow-up survey focused specifically on cognitive symptoms and their functional impact. A total of 306 participants completed this second survey, all of whom were a subset of the original study cohort. To our knowledge, this represents the first systematic real-world assessment of cognitive manifestations and their functional impact in individuals with TMAU.

Among the 306 respondents, brain fog and related cognitive difficulties were both frequent and functionally consequential (Table 6). Young adulthood (18-30 years) was the most frequently reported age at first onset (n = 141, 46.1%), followed by adolescence (n = 52, 17.0%) and childhood (n = 35, 11.4%). Twenty-two respondents (7.2%) selected the combined "never experienced brain fog/cognitive symptoms / not applicable" category.

**Table 6.**
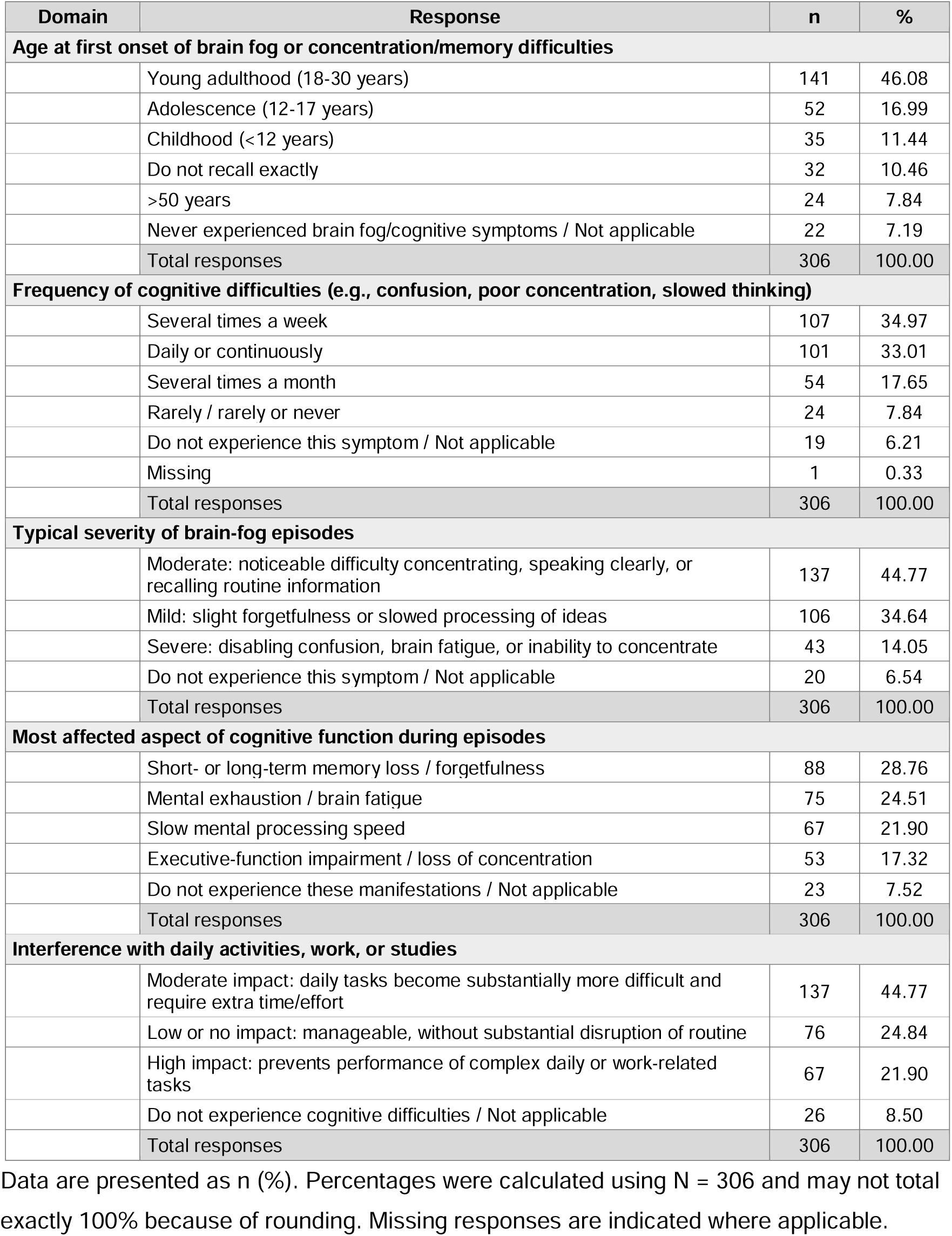
Cognitive manifestations and functional impact of brain fog associated with TMAU (N = 306)

Cognitive difficulties were frequently recurrent: 107 respondents (35.0%) experienced them several times per week and 101 (33.0%) daily or continuously, together accounting for 68.0% of the cognitive-survey cohort. Typical episodes were rated as moderate by 137 respondents (44.8%) and severe by 43 (14.1%). The most commonly affected cognitive domain was short- or long-term memory loss or forgetfulness (n = 88, 28.8%), followed by mental exhaustion or brain fatigue (n = 75, 24.5%), slowed mental processing (n = 67, 21.9%), and executive-function impairment or loss of concentration (n = 53, 17.3%).

Functional consequences were also substantial. Moderate interference with daily activities, work, or studies was reported by 137 respondents (44.8%), while 67 (21.9%) reported high interference that prevented performance of complex daily or work-related tasks. Overall, 204 of 306 respondents (66.7%) reported at least moderate functional disruption. Importantly, greater frequency of cognitive symptoms was significantly associated with greater symptom severity (Spearman’s ρ = 0.451, p < 0.001; Figure 1).

**Figure 1.**
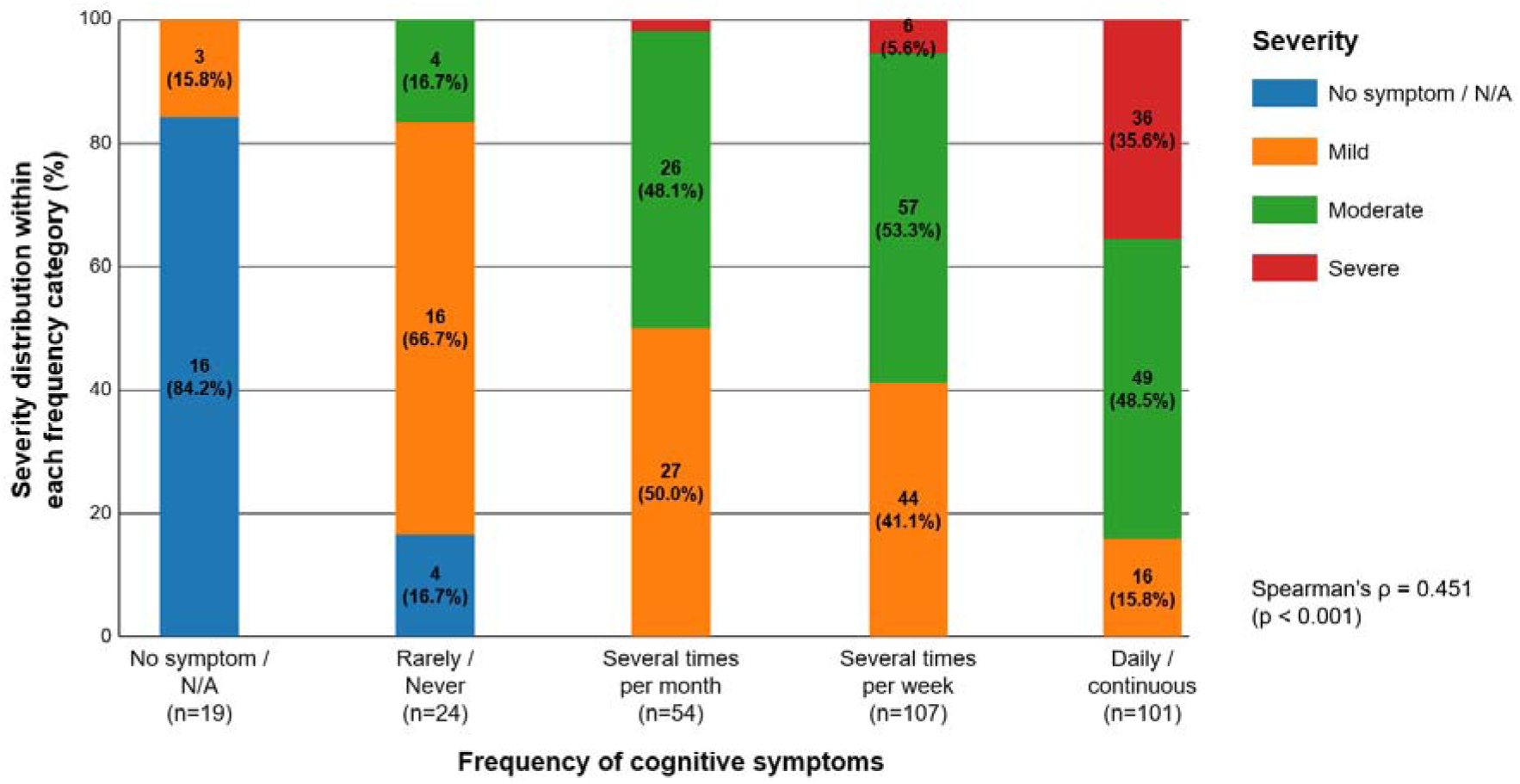
Association between frequency and severity of cognitive symptoms in individuals with TMAU. Severity distributions are shown as percentages within each cognitive-symptom frequency category, with counts and percentages indicated within each bar. Among participants reporting cognitive symptoms with complete paired frequency and severity data (n = 282), greater symptom frequency was significantly associated with greater severity (Spearman’s ρ = 0.451, p < 0.001). Participants reporting no cognitive symptoms or not-applicable responses were excluded from the correlation analysis.

## Discussion

This multinational survey demonstrates that the burden of TMAU extends well beyond malodor, affecting psychological well-being, education, employment, social participation, healthcare experiences, and perceived adequacy of treatment. These findings are consistent with previous qualitative and survey-based studies describing stigma, anxiety, social withdrawal, and impaired daily functioning [9, 10], while extending this evidence to a larger and geographically diverse cohort.

Puberty and adolescence may represent a particularly vulnerable period in TMAU. Hormonal changes can exacerbate the metabolic phenotype [3, 11, 12], while adolescence is simultaneously characterized by heightened sensitivity to peer evaluation, identity, and social belonging. Evidence from other chronic conditions indicates that illness-related stigma and bullying during this developmental period can adversely affect psychological adjustment [13, 14]. The convergence of biological symptom fluctuation and social vulnerability may therefore contribute to the substantial life-course burden reported by participants.

The effects of TMAU can extend beyond embarrassment and interfere with school, work, and social participation. Because patients often appear otherwise healthy, others may not recognize the odor as a symptom of a metabolic disorder. The odor can also be intermittent and may not be present during a medical visit, which can make the condition difficult to understand or verify [7]. As a result, patients may be judged as having poor hygiene or may experience rejection and avoidance from others. Greater awareness of TMAU among healthcare professionals, schools, and workplaces may help reduce these misunderstandings and support appropriate accommodations.

Delayed or absent recognition remains a major challenge. Intermittent symptoms, limited clinician familiarity, and restricted access to biochemical testing can complicate diagnosis [4, 11, 15, 16]. Because olfactory assessment alone is unreliable, urinary TMA/TMAO analysis and, where appropriate, genetic testing remain central to evaluation [11, 12, 16]. The broader rare-disease literature similarly documents prolonged diagnostic journeys, repeated consultations, and psychosocial consequences of diagnostic uncertainty, especially when symptoms begin in childhood or adolescence [17–19]. Together, these findings argue for clearer referral pathways and greater access to appropriate biochemical and genetic evaluation. They also underscore the importance of distinguishing confirmed TMAU from other causes of body or breath malodor rather than assuming that all self-reported odor symptoms represent the same underlying disorder.

Current management relies largely on dietary modification, hygiene measures, and other approaches intended to reduce TMA production or excretion, but treatment responses remain variable [20, 21]. Dietary intervention itself may be burdensome and requires nutritional supervision, particularly because choline is an essential nutrient [22, 23]. Future therapies should therefore be evaluated not only through biochemical endpoints but also through outcomes meaningful to patients, including social participation, emotional well-being, sleep, daily functioning, and cognitive symptoms [24].

Because TMA is generated primarily through microbial metabolism of dietary precursors in the intestine, the gut microbiome represents an attractive upstream therapeutic target in TMAU. Experimental studies have shown that selective inhibition of microbial TMA-producing pathways can substantially reduce intestinal TMA generation without broadly suppressing the microbiota, supporting the feasibility of mechanism-based microbiome modulation [25, 26]. Other preclinical studies have further suggested that selected microbial strains may directly reduce intestinal TMA levels, although such approaches remain investigational. Thus, strategies that inhibit microbial TMA production or introduce microorganisms capable of degrading TMA within the intestinal lumen could potentially reduce systemic TMA exposure before absorption. This concept is particularly relevant to TMAU, in which the host capacity to metabolize TMA is intrinsically impaired, and warrants further evaluation as a microbiome-directed therapeutic approach.

The cognitive findings identify a previously underrecognized dimension of TMAU, with patients reporting brain fog [27, 28], memory difficulty, impaired concentration, slowed thinking, and mental fatigue. Importantly, the observed association between symptom frequency and severity suggests that these complaints may represent a clinically meaningful component of the patient-reported burden. Their underlying mechanism, however, remains uncertain. Psychological distress, sleep disturbance, chronic social hypervigilance, and restrictive dietary practices may all contribute to perceived cognitive dysfunction. Although several studies have implicated elevated TMAO in neuroinflammation and cognitive impairment [29–31], this mechanism may not be directly applicable to TMAU patients, in which impaired FMO3 activity limits the conversion of TMA to TMAO and instead results in accumulation of TMA. Notably, Hoyles et al. reported that physiologically relevant concentrations of TMA impaired blood–brain barrier function and disrupted tight-junction integrity [32]. These findings raise the possibility that TMA itself, rather than TMAO, could have previously underrecognized effects on the cerebrovascular interface in TMAU. Whether elevated TMA contributes to the cognitive manifestations observed in individuals with TMAU remains unknown and warrants further mechanistic investigation.

Another plausible contributor is prolonged dietary choline restriction. Restriction of choline-rich foods has historically been used to reduce intestinal TMA production and malodor in TMAU, although adequate choline intake remains important because choline is an essential nutrient [20, 22]. Choline also serves as a precursor for acetylcholine, a neurotransmitter with important roles in attention, learning, and memory [33]. Experimental studies have shown that prolonged dietary choline deficiency can reduce acetylcholine synthesis and release, particularly in the hippocampus, and impair memory performance [33, 34]. Thus, excessive or prolonged choline restriction could theoretically contribute to cognitive complaints in some individuals with TMAU through reduced cholinergic substrate availability. This possibility remains speculative, as dietary choline intake and cholinergic function were not assessed in the present study, and direct evidence linking dietary choline restriction to cognitive impairment in TMAU is currently lacking. Nevertheless, this hypothesis underscores the importance of maintaining nutritionally adequate choline intake and evaluating dietary status in future studies of cognitive function in TMAU.

A major strength of this study is the breadth of patient-reported information collected across a large multinational cohort, enabling multiple dimensions of TMAU-related burden to be examined together. Several limitations should nevertheless be considered. The cross-sectional design precludes causal inference, and reports of childhood and adolescent experiences were largely retrospective because most participants were adults, introducing potential recall bias. Recruitment through online patient communities may also have resulted in self-selection bias, while uneven geographic representation limits cross-cultural comparisons. In addition, diagnostic status was heterogeneous, with many participants lacking biochemical or genetic confirmation; therefore, the findings should be interpreted as real-world patient-reported experiences rather than prevalence estimates from a uniformly confirmed TMAU population. Finally, several items permitted multiple responses, and these percentages represent non-mutually-exclusive selections. Future prospective studies should include better-characterized cohorts and validated assessments of cognitive, psychiatric, and quality-of-life outcomes.

## Conclusion

TMAU should not be viewed solely as a disorder of malodor. Our findings demonstrate a broader burden involving psychosocial distress, disruption of daily functioning, and previously underrecognized cognitive manifestations. Earlier diagnosis, multidisciplinary support, and development of mechanism-based therapies should therefore be accompanied by patient-centered outcomes, including cognitive function and quality of life.

## Supporting information

Supplementary Testimony

## Data Availability

The datasets generated and analyzed during the current study are available from the corresponding author on reasonable request, subject to patient privacy and data-protection protocols.

## Declarations

### Ethics approval and consent to participate

The study protocol and informed-consent procedures were reviewed and approved by the Latin American Patients’ Academy (ALAPA). Electronic informed consent was obtained from all adult participants. For participants under 18 years of age, informed consent was obtained from their parents or legal guardians, along with the participants’ own assent.

### Consent for publication

Not applicable. No individual personal details, images, or identifiable videos are included in this manuscript.

## Authors’ contributions

LAGL conceptualized and designed the study, developed the psychological evaluation instruments and technical architecture on KoboToolbox, led the primary data collection and curation, and drafted the initial manuscript. SSY supervised the overall project, contributed to the survey module design and methodological framework, provided resources, and critically reviewed and revised the manuscript. VME contributed to participant recruitment, survey dissemination, and secondary data synthesis. CVW reviewed and structured the bioethical framework and informed consent protocols. All authors read and approved the final manuscript.

## Acknowledgements

We express our deepest gratitude to the TMAU patient community, whose trust, courage, and participation made this study possible and helped elucidate the psychosocial realities of living with a rare condition. We sincerely thank our clinical psychology colleagues, Pedro Alonso Forero Saboyá, Violeta Violet, Verónica Pinatti, and Carolina Oliveto for their expert review on item sensitivity and ethical rigor. We also acknowledge Dr. Tania Alejandra Sapuyes Chávez, Ph.D. of Fundación Universitaria del Área Andina for her academic mentorship and institutional support.

