## Supplementary Testimony for "Beyond Malodor: A Multinational Patient-Reported Study of the Diagnostic, Psychosocial, and Cognitive Burden of Trimethylaminuria"

**Supplementary Information**

**Additional file 1**

**Qualitative Patient Narratives on the Psychosocial Impact of TMAU**

**Scope and presentation**

**The following participant-reported narratives are provided as qualitative supplementary material to complement the survey findings presented in the main manuscript. They illustrate the lived psychosocial impact of the condition across education, employment, social participation, relationships, daily activities, and mental well-being. Responses are presented in English; translated responses are identified where indicated in the source material.**

-------------------------------------------------------------------------------------------------------------------------- Potentially identifying details, including precise ages and selected family or medical details, have been generalized or omitted to protect participant confidentiality; wording may therefore differ slightly from the original responses.

1. I took antibiotics, and then all of this started; my life changed. Every time I go out, I feel terrible; the mocking and rejection cause my depression and suicidal thoughts to persist.

2. I only attended elementary school in person; middle school, high school, and university were completed via distance learning, which affected my verbal development. Additionally, I did not finish university because, sadly, I thought it was pointless if I would never be able to practice my profession.

3. When traveling by public transportation, I experience anxiety, which triggers heavy sweating and desperation, releasing an intense odor of feces, sulfur, or something very similar to gunpowder, coupled with a strong smell of urine. This causes me to isolate myself, always being alone and locked away with no desire to go out due to my overwhelming fear of people's reactions; I have even been on the verge of suicide.

4. I have many terrible experiences locked in my mind that have caused me a great deal of stress. Among them, while traveling, everyone was staring at me and I heard comments like "it smells like a corpse" and worse; I hid in a bathroom and ultimately could not continue my journey. This led to a severe conflict with someone close to me; I panicked, sweated profusely, and felt like my heart was going to jump out of my chest.

At school as a young girl, nobody wanted to do homework with me; when they invited others to their houses, they never invited me. I would hear them say I smelled like fish and that "she needs to shower." Throughout my life, I have only maintained two sincere friendships because the others drifted away, and even then, I know these two friends do not introduce me to their other social circles.

In close relationships, I have experienced rejection; some people were even capable of telling me my breath stank. I have also endured a lot of physical and verbal abuse, and I tolerate this because I feel I do not deserve anyone else and that no one else will ever look at me. Also, due to financial constraints, I cannot work; I am terrified of going out there to work because I already went through that. I abandoned a public-facing career because I couldn't concentrate due to comments like "it smells like trash."

My life in general has been highly chaotic; even family members would tell me to go bathe right after I had just showered. I also remember one day it took me over two hours to get home from work, even though it wasn't far, because I kept getting on and off the bus to avoid being with the same people, or I would just walk. I took another course, but with immense effort, as my classmates refused to do practical training with me. At family gatherings, I hear children make humiliating comments about my odor; with everything I have to hear whispered during those gatherings, I leave with my mental health completely shattered, and I promised myself I will never go back. Consequently, I isolate myself more and more every day.

During the lockdown, for me, it was the best thing that could have happened; I know that sounds very extreme, but having no contact with people was the best thing, it gave my mind a rest for a while. Even so, neighbors would complain saying I didn't throw out my trash. I have so many traumatic episodes in my mind that it is hard for me to talk about them, and I would never finish. I was locked up without leaving my house for a long time; now, someone close to me has given me a little strength to keep living, because I really have thought about ending it all just to find peace.

I also recall a terrible experience during a trip. When I went to a shared dining area, many people would stand up and leave as soon as I entered and stared at me judgmentally. One person walked past me and said "she's so filthy." I spent the rest of the trip isolated and enjoyed nothing; someone traveling with me had to bring food to me, which generated conflicts. During those past trips, my illness did not have a name yet, but I knew something was wrong with me. Right now, I no longer travel unless it is by car, and I only do it when necessary for family, but the only safe place is my home.

I am so tired of people gifting me personal hygiene products; one day a family member told me, "here, take this so you can clean yourself and smell nice," but it was said with sheer malice. I cried for a week, and another family member just said "it was a joke", it is always like that. I now have a phobia of going outside; my self-esteem is on the floor. I want to pass unnoticed, so I no longer dress up like I used to in order to avoid drawing attention. At a school-related meeting, people would open the window as soon as I arrived, and one person walked out saying "I can't stand this smell," while everyone stared at me as they left.

It is a constant, crushing weight in your head; being known in your entire residential area for this, walking by and having people start whispering, the pain we carry inside and the constant tightness in our chest is immense. Thank you for listening to us, thank you for believing in us; please keep helping us because the only relief I used to see was in passing away, and now I have hope. I want my family to be happy and to experience something closer to a normal life, and although no one can give us back the time we lost, at least let our final years be dignified. I want to stay motivated, but I am terrified that it won't work; I usually try to be positive, but this medical condition has made me pessimistic, unable to believe in much, and I know that is not who I am because I have lost my personality too. Thank you.

5. I basically had to change my entire life. I had to change my career. I cannot work in person, and I need to isolate myself at home 24 hours a day, 7 days a week.

6. The most severe impact of my condition is the professional hardship I am currently facing. I live in constant anguish because I am on the verge of being fired due to my condition; my coworkers report me because of my odor, and navigating this situation is extremely difficult for me because I have significant family financial responsibilities and cannot afford to lose my job.

7. It stripped away my self-worth; I have never felt important or like I am worth anything to another person. I couldn't pursue a career because even the professors looked down on me. Since I was a very young girl, I have tried to take my own life, and it remains a constant thought.

8. I have stopped participating in many family activities because of this condition; we no longer go on vacations because I cannot bring myself to board an airplane or a public bus. This condition completely transformed my life; I have become a solitary person, isolated from the world, suffering from panic and anxiety attacks.

9. Something that deeply scarred me occurred during my adolescence: when they were discussing hygiene at school, everyone turned around to look at me and laugh at me. :( Since then, I decided never to return to that school; I lost the school year, and it severely affected me emotionally. I no longer wanted to socialize with anyone; people have treated me horribly because of this condition.

10. There were many situations, the most uncomfortable and difficult being in middle school, where I was subjected to constant mocking and hurtful comments. Also in enclosed spaces like movie theaters, among others.

11. Always alone. Every time I had my menstrual period, it was an absolute nightmare. I abandoned my studies, and I no longer know how to interact with people.

12. *[Translated from Italian]* The greatest distress occurs in the workplace. As soon as I was able to, I resigned from my job.

13. It completely changed my life; I have been rejected by schools, and I have noticed rejection and comments since elementary school. It has also been very difficult to face this in adulthood, as I receive frequent comments at my university, which has prevented me from attending classes on many occasions.

14. I love going to school and actively participating in my community; however, this condition made me withdraw and become afraid of speaking to people.

15. It is like living in a nightmare. You cannot leave the house without being pointed at or ridiculed. It is an omnipresent suffering that has no end, and I feel like I have no future.

16. All of this started when I took antibiotics. The worst part is that it is causing me depression and suicidal ideation. It damages you mentally.

17. I received death threats and had to flee my home; I couldn't finish my technical degree, I was only missing my practical internship.

18. My most difficult struggle is seeing my condition reflected in a close family member. I have faced many challenging situations related to the odor in the workplace, but what devastates me completely is that someone close to me also has to suffer from this horrible disease.

19. This condition makes daily life incredibly difficult. It is not only hard to expose and socialize ourselves, but it also damages relationships with neighbors, family members, colleagues, etc. It affects us both emotionally and psychologically.

20. My life ceased to be a life from day one when this illness awakened in my body. Society forces me to completely isolate myself from the world; I am unable to even go to a local store for essential items. My social circle distanced themselves from me, and the hardest part is securing a job. Nervousness, stress, depression, and suicidal thoughts are my daily bread.

21. My life is entirely bound to this condition. I do not use public transportation, I do not dare to converse with strangers, I experience daily anxiety, and I have suffered from depressive episodes.

22. It has affected me so deeply that even my family is suffering, because I have already attempted to end my life three times because nobody will come near me due to my odor.

23. I am a suicide survivor; I shouldn't be alive. Human cruelty, discrimination, medical rejection, the apathy of my family, and the lack of understanding of what was happening to me broke me.

24. Since middle school, I felt rejected by people; almost no one spoke to me because they thought I was dirty. I have lived with rejection and bullying since early adolescence (age range 11–15), and I am currently in the 46–50 age range. Because of this, I am extremely shy, have low self-esteem, and suffer from suicidal thoughts.

25. The hardest part was having to complete almost my entire education online (except for elementary school) and dropping out halfway through university because of the persistent feeling that I would never be able to practice my profession anyway.

26. My symptoms began when I was in the 16–20 age range, and I remember being terrified of going to school. As my symptoms worsened nearing college, I decided to major in something I thought would keep me isolated and in the background. College came, and I had to stop my studies because of this condition. I returned, and then the pandemic disrupted my studies. I decided to finish my degree online, and by the time I was in the 21–25 age range, I had no hope. I didn't think I would make it. I hoped I wouldn't have to live another day with this condition. I worry about my future and the future of those around me who share this burden with me. Even if this doesn't help me, I hope it at least brings comfort and hope to those who also feel alone.

27. This has caused me to isolate myself completely. I have no desire to leave my house; only inside my home do I feel safe. I suffer from social anxiety and suicidal thoughts. It is the most humiliating thing that has ever happened to me.

28. Everything began after an extended course of antibiotics when I was in the 16–20 age range. Living has become an absolute challenge because multiple factors exacerbate the odor—starting with diet, weather changes, and even menstruation. Consequently, all I have received over the years are mockery, hostile attitudes, judgmental stares, and harsh words. For these reasons, I only go out to work and avoid any other type of outing. Thank you for reading me. :)

29. Without a doubt, the most difficult period was during my high school years. My classmates called me "fish" because of the odor associated with my condition. I was also mocked during university. Despite these painful experiences, I managed to achieve almost all of my academic and personal goals. However, these situations had a major impact on my emotional well-being and the way I related to others.
